# RESISTANCE CONFERRING MUTATIONS IN SARS-CoV-2 DELTA FOLLOWING SOTROVIMAB INFUSION

**DOI:** 10.1101/2021.12.18.21267628

**Authors:** Rebecca J Rockett, Kerri Basile, Susan Maddocks, Winkie Fong, Jessica E Agius, Jessica Johnson Mackinnon, Alicia Arnott, Shona Chandra, Mailie Gall, Jenny Draper, Elena Martinez, Eby M Sim, Clement Lee, Christine Ngo, Marc Ramsperger, Andrew N Ginn, Qinning Wang, Michael Fennell, Danny Ko, H Ling Lim, Nicky Gilroy, Matthew V N O’Sullivan, Sharon C-A Chen, Jen Kok, Dominic E Dwyer, Vitali Sintchenko

## Abstract

Several Severe Acute Respiratory Syndrome Coronavirus-2 (SARS-CoV-2) neutralising monoclonal antibodies (mAbs) have received emergency use authorisation by regulatory agencies for treatment and prevention of Coronavirus Disease 2019 (COVID-19), including in patients at risk for progression to severe disease. Here we report the persistence of viable SARS-CoV-2 in patients treated with sotrovimab and the rapid development of spike gene mutations that have been shown to confer high level resistance to sotrovimab *in vitro*. We highlight the need for SARS-CoV-2 genomic surveillance in at risk individuals to inform stewardship of mAbs use and prevent potential treatment failures.

## INTRODUCTION

Severe Acute Respiratory Syndrome Coronavirus 2 (SARS-CoV-2) which causes Coronavirus disease 2019 (COVID-19) has spread rapidly worldwide causing over 258 million infections and 5 million deaths^1^, prompting the development of vaccines and antiviral agents at an unprecedented pace.^2^ Several SARS-CoV-2-neutralizing monoclonal antibodies (mAbs) have been developed and received emergency use authorisation by regulatory agencies with additional mAbs currently advancing through phase 3 clinical trials. These therapeutics have been registered for treatment of mild to moderate COVID-19 disease in those at risk of progressing to severe disease. These agents target the SARS-CoV-2 spike (S) glycoprotein which consists of two functional subunits: S_1_ which includes the receptor-binding domain (RBD) and N-terminal domain (NTD) and mediates host attachment, and S_2_ subunit responsible for fusion of the virus and cellular membranes. The emergence of SARS-CoV-2 variants of concern (VOC) carrying mutations and deletions in the RBD, NTD and S_2_ subunit has highlighted the need for a more targeted utilization of these advanced therapeutics. Several mAbs cocktails (e.g., bamlanivimab/etesevimab) appear to be have reduced activity against viruses with the E484K and K417N/T mutations found in the lineages B.1.525, B.1.526 and VOC.^3^ Other mAbs, such as sotrovimab (VIR-7831, GlaxoSmithKline, Australia Pty Ltd), target more conserved viral epitopes.

Sotrovimab is a human engineered monoclonal antibody that neutralizes SARS-CoV-2 and other sarbecoviruses, including SARS-CoV-1.^4^ Sotrovimab acts by binding to a conserved epitope within the RBD, resulting in virus neutralization. The conservation of the epitope targeted by sotrovimab is supported by preservation of its activity *in vitro* against SARS-CoV2 VOC Alpha, Beta and Delta; however sotrovimab effectiveness is yet to be determined for the Omicron VOC.^5^ Sotrovimab’s safety and efficacy was evaluated in the phase 3, multicenter, randomised, double-blind, placebo-controlled COMET-ICE (COVID-19 Monoclonal Antibody Efficacy Trial–Intent to Care Early) trial. Trial recruitment targeted high-risk adults with symptomatic COVID-19 and interim results demonstrated a reduction in the risk of hospitalization (for >24 hours) or death from 7% in the placebo group to 1% in the sotrovimab group (85% relative risk reduction). Whilst evidence of sotrovimab effectiveness to prevent severe COVID-19 led to its approval for emergency use in the US, Singapore, Europe and Canada, Australia was one of the first countries to issue formal regulatory approval.^6–8^ However, the use of SARS-CoV-2-specific monoclonal antibodies targeting a single viral epitope warrants caution as *in vitro* ^9–11^ and clinical case studies have demonstrated rapid development of mutations conferring resistance after exposure to various SARS-CoV-2 mAbs.^12–17^ Indeed, the acquisition of mutations in the sotrovimab target epitope at S protein amino acid positions 335-361 was reported during COMET-ICE.^4^ Phenotypic characterization of these mutations demonstrated that high-level sotrovimab resistance (100 to 295-fold reduction in neutralization) was conferred by E340K/A/V, while a 192 to 304-fold reduction was also noted with the appearance of mutation P337L/K/R. ^12^ The acquisition of consensus mutations at S:E340K/A was also documented for 4 of 45 participants within the COMET-ICE trial. Observations of induced *in vitro* and clinical resistance has prompted the development of mAbs cocktails that simultaneously target multiple SARS-CoV-2 epitopes.^13^ Here we report the rapid development of mutations *in vivo* that have been shown to confer high level resistance to sotrovimab *in vitro* and present a template for genomics guided use of mAbs.

## METHODS

### Patient cohort

A total of 100 patients received sotrovimab between 22^nd^ August 2021 and 13^th^ November 2021 at a single center in Australia. Sotrovimab was provisionally registered for use by the Australian Therapeutics Goods Administration in August 2021, with limited supply. Treatment, a single 500mg infusion, was targeted at patients within five days of symptom onset who presented with risk factors for progression to severe disease.^4^

Of the 100 patients that received sotrovimab, 23 had persistent SARS-CoV-2 RNA detected by reverse transcriptase – real time polymerase chain reaction (RT-PCR) for more than 10 days post-infusion (Supplementary Figure S1). A total of 68 patients did not have a follow-up SARS-CoV-2 RT-PCR test following treatment. Longitudinally-collected respiratory tract specimens pre- and post-sotrovimab were available for eight patients (35%) of the persistently RT-PCR positive patients and were thus investigated in this study (R001-R008).

Case demographic and clinical (co-morbidities, treatment and COVID-19 vaccine status) information was compiled for the study cohort (Table 1, Supplementary Table S1). Complete COVID-19 vaccination was defined as two doses of BNT162b2 (Comirnaty, Pfizer/BioNTech) received at least 7 days before testing positive to SARS-CoV-2 and in accordance with COVID-19 local vaccination recommendations.^14^ Partial vaccination was defined as either one dose of vaccine or receipt of the second dose <7 days from SARS-CoV-2 RNA detection by RT-PCR. Cases R009 and R010 were identified as asymptomatic household contacts of study cohort case R001 and were also included in the study cohort (Supplementary Table S1). Ethical and governance approval for the study was granted by the Western Sydney Local Health District Human Research Ethics Committee (2020/ETH02426) and (2020/ETH00786)

**Table 1.** Characteristics of cases treated with sotrovimab and requiring hospitalization

| **Case** | **Age (10 year range)** | **Vaccination Status** | **Co-morbidities and risk factors for severe COVID-19** | **Hospital admission** | **Other antiviral therapy** | **Spike mutations detected following Sotrovimab therapy** |
| --- | --- | --- | --- | --- | --- | --- |
| **R001** | 30-40 | Complete (Pfizer/BioNTech) | Lung Tx | Ward | Remdesivir | S:E340K<br>S:E340V |
| **R002** | 30-40 | Partial (Pfizer/BioNTech) | CVID | ICU - ECMO, | Sarilumab, Remdesivir | S:E340A<br>S:P337L |
| **R003** | 20-30 | Partial (Pfizer/BioNTech) | Renal Tx<br>CMV<br>BK viraemia | Ward |  | S:E340K |
| **R004** | 70-80 | Not vaccinated | Myeloproliferative disorder† | Ward |  | S:E340K<br>S:E340A<br>S:E340V |
| **R005** | 70-80 | Not vaccinated | Interstitial lung disease† | N |  | Not detected |
| **R006** | 50-60 | Not vaccinated | Renal Tx, MM, T2 DM | Ward | Remdesivir | Not detected |
| **R007** | 60-70 | Partial (Pfizer/BioNTech) | ESRF on HD/T2DM | Ward | Sotrovimab | Not detected |
| **R008** | 70-80 | Complete<br>(Pfizer/BioNTech) | CLL, ESRF on HD, T2 DM | Ward |  | Not detected |

**Key:** CLL – chronic lymphocytic leukaemia treated with intragram; Complete - two doses of BNT162b2 (Comirnaty, Pfizer/BioNTech) received 7 days before testing positive to SARS-CoV-2 in accordance with COVID-19 local vaccination recommendations<sup>14</sup>; CMV – cytomegalovirus; CVID – Common variable Immunodeficiency, receives weekly IVIg; ECMO – extracorporeal membrane oxygenation, without intubation; ESRF- end stage renal failure; F – female; ICU – intensive care unit; IVIg- intravenous immunoglobulin; M – male; MM – multiple myeloma; N – No; Partial – either one dose of vaccine or receipt of the second dose <7 days from SARS-CoV-2 RNA detection by RT-PCR; T2 DM – Type 2 Diabetes Mellitus; Tx – transplant; Y – yes; † - passed away

### SARS-CoV-2 culture

Respiratory tract specimens that had detectable SARS-CoV-2 RNA by RT-PCR were cultured in vero E6 cells expressing transmembrane serine protease 2 (VeroE6/TMPRSS2; JCRB1819) as previously outlined (Supplementary Figure S2).^15^ Briefly, cell cultures were seeded at 1-3×10^4^ cells/cm^2^ in Dulbecco’s minimal essential medium (DMEM, Lonza, Basel, Switzerland) supplemented with 9% foetal bovine serum (FBS, HyClone). Media was replaced within 12 hours with inoculation media containing 1% FBS with the addition of penicillin (10,000 U/mL), streptomycin (10,000 μg/mL) and amphotericin B deoxycholate (25 μg/mL) (Lonza) to prevent microbial overgrowth and then inoculated with 100-500 μL of SARS-CoV-2 positive respiratory sample. The inoculated cultures were incubated at 37°C in 5% CO_2_ for 4 days and observed daily for cytopathic effect (CPE). Routine mycoplasma testing using RT-PCR was performed to exclude cell line mycoplasma contamination and culture work was undertaken under physical containment laboratory level 3 (PC3) biosafety conditions. The presence of CPE and increasing viral load was indicative of positive SARS-CoV-2 culture. Culture supernatant was harvested four days after inoculation and stored at -80°C.

### SARS-CoV-2 metagenomics

RNA extracts from case R004 (day 2, 6, 11 and 15) were rRNA depleted using the NEBNextÒ rRNA Depletion Kit (Human/Mouse/Rat) (New England Biolabs, Notting Hill, Australia). The rRNA-depleted RNA was then converted to cDNA using LunaScriptÒ RT SuperMix Kit (New England Biolabs) and the second strand was synthesised with NEBNextÒ Ultra II Non-Direction RNA Second Strand Synthesis Module (New England Biolabs). Libraries were prepared in duplicate using Nextera XT (Illumina, Melbourne, Australia) according to manufacturer’s instructions and sequenced with paired end 100 bp chemistry on the NextSeq 2000 (Illumina).

### SARS-CoV-2 genomics

Tiling PCR was used to amplify the entire SARS-CoV-2 genome from RNA extracts of clinical specimens using primers outlined in the Midnight sequencing protocol.^16^ Each PCR included 12.5μL Q5 High Fidelity 2x Master Mix (New England Biolabs), 1.1μL of either pool 1 or pool 2 10μM primer master mix, 2.5μL of template RNA and molecular grade water was added to generate a total volume of 25μL. Cycling conditions were initial denaturation at 95°C for 2 min, then 35 cycles of: 95°C for 30s, 65°C for 2 min 45s, and a final extension step of 75°C for 10 min. Pool 1 and pool 2 amplicons were combined and purified with a 1:1 ratio of AMPureXP beads (Beckman Coulter) and eluted in 30μL of RNAase free water. Purified products were quantified using Qubit™ 1x dsDNA HS Assay Kit (Thermo Fisher Scientific) and diluted to the desired input concentration for library preparation. Sequencing libraries were prepared using Nextera XT (Illumina) according to the manufacturer’s respective instructions and pooled with the aim of producing 1×10^6^ reads per library. Sequencing libraries were then sequenced with paired end 76 bp chemistry on the iSeq or MiniSeq (Illumina) platforms.

### Bioinformatic analysis

Raw sequence data were processed using an in-house quality control procedure prior to further analysis as described previously.^17,18^ De-multiplexed reads were quality trimmed using Trimmomatic v0.36 (sliding window of 4, minimum read quality score of 20, leading/trailing quality of 5 and minimum length of 36 after trimming).^19^ Briefly, reads were mapped to the reference SARS-CoV-2 genome (NCBI GenBank accession MN908947.3) using Burrows-Wheeler Aligner (BWA)-mem version 0.7.17^20^, with unmapped reads discarded. Average genome coverage was estimated by determining the number of missing bases (Ns) in each sequenced genome. Variant calling and the generation of consensus sequences was conducted using iVar^21^, with soft clipping over primer regions (version 1.2.1, min. read depth >10x, quality >20, min frequency threshold of 0.1). Single nucleotide polymorphisms (SNP) were defined based on an alternative frequency >0.75 whereas Minority allele Frequency Variants (MFV) were defined by an alternative frequency between 0.1 and 0.75. Variants falling in the 5’ and 3’UTR regions were excluded. Polymorphic sites that have previously been highlighted as problematic were monitored.^22^ To ensure the accuracy of variant calls only high-quality genomes with >90% genome coverage and a mean depth of >1000x were included. The MFV calls were excluded in the base pair either side of the 5’ or 3’-end of indels due to potential mis-mapping. SARS-CoV-2 lineages were inferred using Phylogenetic Assignment of Named Global Outbreak LINeages v1.2.86 (PANGO and PLEARN).^23,24^ Representative SARS-CoV-2 genomes collected between July to November 2021 (n=1,300, ≥27,000bp in length) were downloaded from Global initiative on sharing all influenza data (GISAID)^25^ EpiCoVÔ, using a global subsampling strategy developed by Nextstrain^26^ Phylogenetic inference and visualisation of the 1,133 high quality consensus SARS-CoV-2 FASTA sequences (GISAID, n = 1,084; study, n = 50) was performed using the Nextstrain conda environment, Augur v13.0.3 (https://github.com/nextstrain/augur), and Auspice (open-source visualisation tool) v2.32.0 (https://github.com/nextstrain/auspice). Wuhan/Hu-1/2019 was used to root the phylogram and Auspice was used to visualise the resulting phylogeny (Supplementary Figure S2). Genomes defined by PANGO as the Delta lineage (n=205) were used to contextualise sotrovimab resistant specimens generated in this study (n=50) (Figure 1B). The GISAID and New South Wales (NSW) genomes were aligned with MAFFT v7.402 (FFT-NS-2, progressive method).^27^ Phylogenetic analysis was performed using the maximum likelihood approach (IQTree v1.6.7 (substitution model: GTR+F+R2) with 1,000 bootstrap replicates.^28^ Graphs were generated using RStudio (version 3.6.1) and phylogenetic trees were constructed using the R package ggtree.^29^

**Figure 1.**
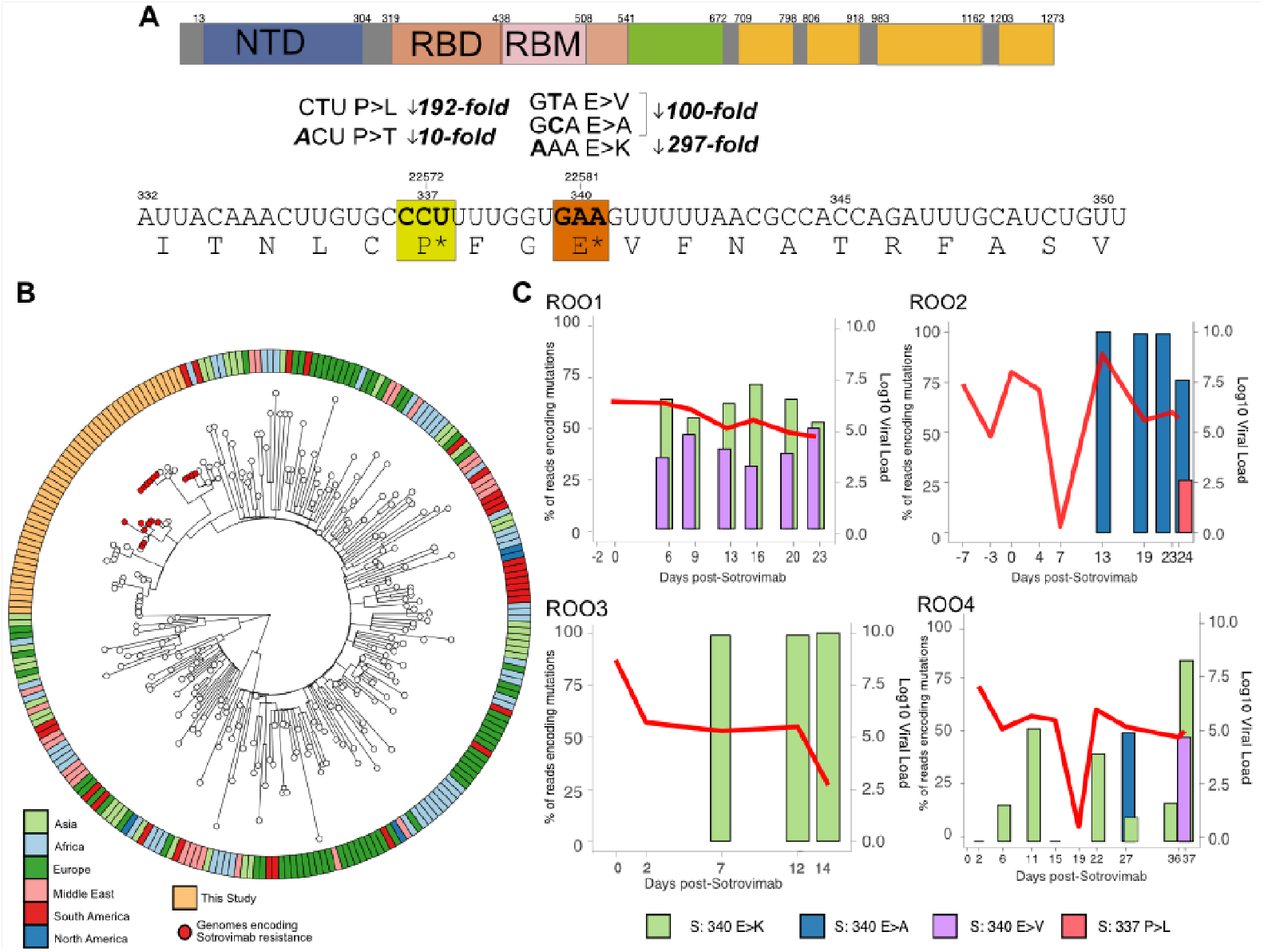
SARS-CoV-2 viral load dynamics and acquisition of resistant mutations following sotrovimab treatment. **Figure 1.** Acquisition of mutations conferring high level resistance to sotrovimab and dynamics of SARS-CoV-2 viral load. A) Depicts the co-ordinates of mutations that are acquired after sotrovimab treatment in the receptor binding domain (RBD) of the spike (S) protein. In this study we uncovered mutations (S:E340K/A/V and S:P337L/T) which have been reported to reduce the susceptibility to sotrovimab by 297, 100, 200, 192 to 10-fold, respectively. B) Subsampled global phylogeny of SARS-CoV-2 VOC Delta, the geographical region of each sequence is indicated in the outer metabar. SARS-CoV-2 genomes sequenced in this study are highlighted in orange in the outer metabar, red nodes indicate genomes that developed resistance-conferring mutations. C) The dynamics of four cases of SARS-CoV-2 infection treated with sotrovimab are outlined on separate graphs; the x-axis indicates the days sampled pre- and post-sotrovimab infusion. The bar graph depicts the acquisition and read frequency (left y-axis) of mutations conferring high levels of resistance to sotrovimab (green S:340E>K, blue S:E340A, purple S:E340V and pink S:P337L) for each case. The overlayed red line graph shows the SARS-CoV-2 viral load at each sampling point (right-hand y-axis scale. All cases were hospitalized during sampling periods.

### Prevalence of S:E340K/A/V and S:P337L mutations

Local and international genomes were interrogated for mutations S:E340K/A/V and S:P337L reported to confer sotrovimab resistance. A total of 11,841 SARS-CoV-2 genomes from NSW, Australia which were collected between 1 June - 11 November 2021 were downloaded from GISAID. Raw sequencing data was available for 17,303 local SARS-CoV-2 genomes generated at the Microbial Genomic Reference Laboratory, New South Wales Health Pathology-Institute of Clinical Pathology and Medical Research (ICPMR). The presence of mutations conferring resistance in viral sub-populations was investigated in variant calling for sequences generated at the ICPMR. International genomes were assessed for the presence of mutations using covSPECTRUM^30^, with all international genomes between 6 January 2020 and 28 November 2021 included (accessed November 28^th^ 2021 https://cov-spectrum.ethz.ch/)

### Respiratory virus detection by RT-PCR

The RNA was extracted using the Viral NA Small volume kit on the MagNA Pure 96 system (Roche Diagnostics GmbH). A previously described RT-PCR^18^, targeting the nucleocapsid gene was employed to estimate the viral load of clinical specimens. A commercially available synthetic RNA control (Wuhan-1 strain, TWIST Biosciences NCBI GenBank accession MN908947.3) was used in 10-fold dilutions starting at 20,000 copies/μL to 2 copies/μL to generate a standard curve and quantify the viral load of each specimen in duplicate.

### Statistical analysis

Mann-Whitney two tailed *t*-tests were used to assess differences in SARSCoV-2 viral load. A significance threshold of <0.01 was used.

## RESULTS

### SARS-CoV-2 viral load before and after treatment

A median viral load of 142,619 copies/μL of SARS-CoV-2 RNA extract (range 2-8.2×10^8^ copies/μl) was detected in longitudinal study cohort specimens collected between seven days pre- and 37 days post-sotrovimab infusion. Longitudinal fluctuations in the viral load were observed for all cohort members (Figure 1B, Supplementary Data S1). No significant difference in SARS-CoV-2 viral load was detected when comparing the viral loads pre- and post-sotrovimab (*p*=0.07508) or between cases with and without resistant mutations after sotrovimab treatment (*p*=0.01046) (Figure 1C, Supplementary Data S1). A substantial drop in viral load was noted in two cases (R004 & R002) that subsequently rebounded after resistance mutations were detected.

### Development of sotrovimab resistance conferring mutations in SARS-CoV-2 populations

Consensus genomes were recovered for 50/59 clinical specimens with median genome coverage and depth of 98.2% and 5,035.5x respectively (sequencing coverage min 92.0% max 99.9%, depth min 1,784.2 max 6,573.0) (Supplementary Data S1). All genomes were found to belong to Pangolin lineage AY.39.1, a sub-lineage of the Delta VOC that currently predominates in Australia and globally (Figure 1A). Specimens from which high-quality SARS-CoV-2 genomes were unable to be obtained had significantly lower viral loads (cohort median Ct 24.79 compared to Ct 34.17 for failed genomic specimens *p*=<0.0000.1) (Figure 1. Supplementary Data S1). Four of the eight patients in this study acquired previously defined RBD mutations between 6-13 days after sotrovimab treatment (Table 1; Figure 1C). All but one case developed the S:E340K mutation which has previously demonstrated the highest resistance to sotrovimab. Read frequencies of S:E340K/A/V mutations generally increased over the course of infection, in two cases (R002 & R003) the proportion of the viral population carrying these mutations exceeded 75% at days 7 and 13, respectively, and remained fixed at subsequent time-points. In cases R002 and R004, S:E340K was initially detected in increasing frequency, subsequently interchanging between S:E340A and S:E340V. In addition, R002 developed a MFV at P337L after fixation of the S:E340K mutation 24 days after sotrovimab. R004 died 37 days after sotrovimab treatment due to non-COVID-19 related underlying conditions. R002 was the only case to receive concurrent treatment with dexamethasone and remdesivir, from day 2 to 6 following sotrovimab. The S:E340K mutation was detected on Day 13 which corresponded to peak viral load for this case. In contrast, a resistance mutation was detected at day 7 for R003 followed by a gradual decline in viral load without additional COVID-19 treatment.

RNA extracted from specimens collected from case R004 underwent confirmatory metagenomic sequencing, which removes bias from SARS-CoV-2 RNA amplification. Between 49 and 577 million reads were generated per specimen which were collected 2, 6, 11 and 15 days after sotrovimab. These reads produced consensus SARS-CoV-2 genomes with >99.9% coverage with 579-55,767X average depth. The results confirmed the presence of the S:E340K mutation six and 11 days after sotrovimab, but was not detected 15 days postinfusion. Household contacts on R001 who became SARS-CoV-2 RT-PCR positive two days after R001, did not harbour mutations that confer resistance to sotrovimab.

### SARS-CoV-2 culture positivity following sotrovimab therapy

All cases that developed mutations conferring sotrovimab resistance had at least one culture-positive specimen after the acquisition of resistant mutations (Supplementary Data S1). Cases R001, R002, R003 and R004 remained culture-positive for 23, 24, 12 and 15 days after sotrovimab treatment, respectively.

### Prevalence of spike RDB E340K/A/V and P337L

Four of the 11841 local SARS-CoV-2 genomes generated in NSW and available on GISAID contained a consensus mutation at position E340K (Supplementary Table S2). Clinical information was only available for two cases, both of which had been treated with sotrovimab. SARS-CoV-2 genomes were generated 5 and 11 days after sotrovimab treatment, respectively.

Resistance mutations S:E340K/A/V and P337L/T/S were detected in the raw variant calling files of an additional six cases. Three cases contained the mutation S:E340K as a consensus mutation in the genome, one case had received sotrovimab 15 days prior, one case had no reported treatment for COVID-19, and clinical information was unavailable for one case. The remaining three genomes contained mutations detected at position P337T/L/S, but no cases had been treated with sotrovimab (Supplementary Table S3).

Of the 527,931 international SAR-CoV-2 genomes 130, 101 and 24 contained the mutation E340K/A/V, respectively. A further 65 genomes contained the mutation S:P337L. The overall prevalence of these mutations was initially very low in March 2020, however the proportion of international sequences carrying all three mutations increased by September 2021 (Supplementary Figures S4-7).

## DISCUSSION

This is the first record of clinical and genomic outcomes following treatment of SARS-CoV-2 infection with sotrovimab and highlights the importance of genomic data to ensure the stewardship of mAbs administration. Prolonged infection with SARS-CoV-2, particularly in immunocompromised individuals, may allow rapid viral adaptations and intra-host variants to emerge. These individuals may receive a combination of new COVID-19 therapies to aid viral clearance and prevent severe disease. As numerous *in vitro* and clinical case studies have highlighted, close monitoring of these infections is warranted to ensure viral clearance and to detect the development of variants that have the ability to evade emerging therapeutic options.^10,31–37^ Our findings demonstrated the acquisition of mutations in the RBD of the SARS-CoV-2 S protein 6-13 days after treatment with sotrovimab. The genomic position of these mutations has previously been suggested to decrease the ability of sotrovimab to neutralize SARS-CoV-2 by 10–297-fold.^4^ The most commonly detected mutation E340K (3 of 4 cases) was reported to confer the highest levels of resistance.

Previous *in vitro* studies have demonstrated that RBD mutations can lead to reductions in the effectiveness of mAbs and natural or vaccine-elicited neutralizing antibodies.^10,34^ This study adds important clinical data to support the experimental evidence. The mutations described were within highly conserved epitopes of SARS-CoV-2 with <250 international genomes reporting S:E340K/A/V changes in the spike protein, and this mutation has not occurred in VOC Alpha, Beta, Gamma and Delta.

Our findings of persistence viral culture positivity in patients developing sotrovimab mutations also has implications for infection control and their release from isolation. Viral culture is used to determine the transmission risk of immunocompromised SARS-CoV-2 positive patients as part of Australian COVID-19 clinical management guidelines.^38^ We document that SARS-CoV-2 was isolated from specimens collected from patients harboring resistant mutations up to 24 days post-sotrovimab treatment. This indicates that individuals may remain infectious after acquiring mutations and therefore can transmit resistant virus. This may result in reduced efficacy of sotrovimab and potentially of other immunotherapies, including vaccination, in sotrovimab-naïve individuals.

Australia has a relatively low incidence of COVID-19 infections and SARS-CoV-2 genomes are sequenced in a high percentage of diagnosed cases. We leveraged on this advantage and screened extensive SARS-CoV-2 genomic data to assess if transmission of these mutations has gone unrecognized. We identified only four genomes available on GISAID (of 11,841) containing the S:E340K mutation and a further six cases harboring the mutations in sub-consensus viral populations. Of these 10 cases, six had received sotrovimab, with no clinical information available for three cases. This indicates that the development of resistant mutations can be unrecognized, but extensive circulation of sotrovimab resistant virus is not evident at this early stage of sotrovimab use. However, most genomic surveillance strategies currently sample a single SARS-Cov-2 genome per diagnosed case, which may not provide sufficient resolution to detect mutations that develop following COVID-19 treatments and to explain the mechanism of breakthrough infections. The recent observation of G339D mutation in spike in the rapidly spreading Omicron VOC is also of concern.^39^ Therefore, the incidence of these mutations in patients treated with sotrovimab may be underestimated.

In conclusion, SARS-CoV-2 genome analysis and culture in patients treated with sotrovimab can assist in monitoring the progress of COVID-19 infection and managing infection control and duration of isolation periods. Post-marketing genomic surveillance of patients that receive monoclonal antibody therapy for SARS-CoV-2 is prudent to minimize the risk of treatment failure, and transmission of potentially more resistant SARS-CoV-2 variants in both healthcare settings and the community.

## Data Availability

Fastq files have been deposited in BioProject PRJNA633948 for all 50 genomes produced in this study. Individual SRA and Global initiative on sharing all influenza data (GISAID) accessions are available in Supplementary Data S1.

## ACKNOWLEDGEMENTS

The authors are grateful to NSW Pathology partner laboratories, ACT Pathology, Douglass Hanly Moir, Australian Clinical Laboratories and Laverty Pathology for referring samples for genomic surveillance. The authors acknowledge the Sydney Informatics Hub and the use of the University of Sydney’s high-performance computing cluster, Artemis. The authors are indebted to all researchers and their organisations who have kindly shared SARS-CoV-2 genome data on GISAID.

## Notes

### Competing Interest Statement

The authors have declared no competing interest.

### Funding Statement

Funding for this study was provided by the Prevention Research Support Program funded by the New South Wales Ministry of Health and New South Wales Health COVID-19 priority funding (round one).

### Author Declarations

Ethical and governance approval for the study was granted by the Western Sydney Local Health District Human Research Ethics Committee (2020/ETH02426)

