## Supplementary material for "RESISTANCE CONFERRING MUTATIONS IN SARS-CoV-2 DELTA FOLLOWING SOTROVIMAB INFUSION"

**Figure S1. Outline of testing details for patients treated with sotrovimab**

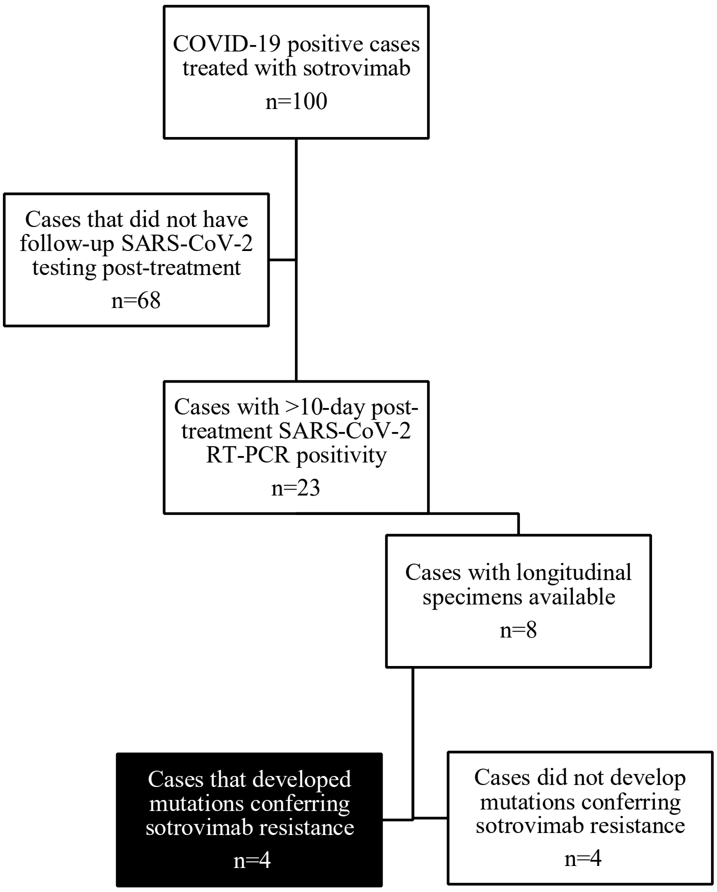


**Key:** COVID-19 – Coronavirus disease 2019 (COVID-19); RT-PCR – reverse transcriptase real time polymerase chain reaction; SARSCoV-2 – Severe Acute Respiratory Syndrome Coronavirus 2

**Table S1. Extended demographic and clinical information**

| Case | Age (yrs) | Co-morbidities | Hospital Admission | Treatment | Vaccination Status |
|---|---|---|---|---|---|
| **R001** | 30-40 | Lung Tx | Y (ward based) | Dexamethasone Remdesivir Sotrovimab | 2 doses of Pfizer/BioNTech, |
| **R002** | 30-40 | CVID | Y, ( ICU - ECMO, without intubation) | Dexamethasone, Sarilumab, Sotrovimab, Remdesivir | 1 dose of Pfizer/BioNTech |
| **R003** | 20-30 | Renal Tx CMV BK viremia | Y, (ward based) | Sotrovimab | 1 dose of Pfizer/BioNTech |
| **R004** | 70-80 | Myeloproliferative disorder † | Y (ward based) | Sotrovimab | Unvaccinated |
| **R005** | 70-80 | Interstitial Lung disease† | Y – (ward based) | Sotrovimab | Unvaccinated |
| **R006** | 50-60 | Renal Tx, MM, T2 DM | Y (ward based) | Baricitinib Dexamethasone Remdesivir Sotrovimab | Unvaccinated |
| **R007** | 60-70 | ESRF/T2DM | Y (ward based) | Dexamethasone Sotrovimab | 1 dose of Pfizer/BioNTech, |
| **R008** | 70-80 | CLL (intragam), ESRF,T2 DM, | Y (ward based) | Dexamethasone Sotrovimab | 2 doses of Pfizer/BioNTech |
| **R009** | 30-40 | Nil known | N | NIL | 2 doses of Pfizer/BioNTech |
| **R010** | 0-10 | Nil known | N | NIL | Unvaccinated |

**Key:** CLL – chronic lymphocytic leukaemia treated with intragram; Complete - two doses of BNT162b2 (Comirnaty, Pfizer/BioNTech) received 7 days before testing positive to SARSCoV-2 in accordance with COVID-19 local vaccination recommendations[14]; CMV – cytomegalovirus; CVID – Common variable Immunodeficiency, receives weekly IVIg; ECMO – extracorporeal membrane oxygenation, without intubation; ESRF- end stage renal failure; F – female; ICU – intensive care unit; IVIg- intravenous immunoglobulin; M – male; MM – multiple myeloma; N – No; Partial – either one dose of vaccine or receipt of the second dose <7 days from SARS-CoV-2 RNA detection by RT-PCR; T2 DM – Type 2 Diabetes Mellitus; Tx – transplant; Y – yes; † - passed away

**Figure S2. Outline of molecular, phenotypic and next generation sequencing methodology**

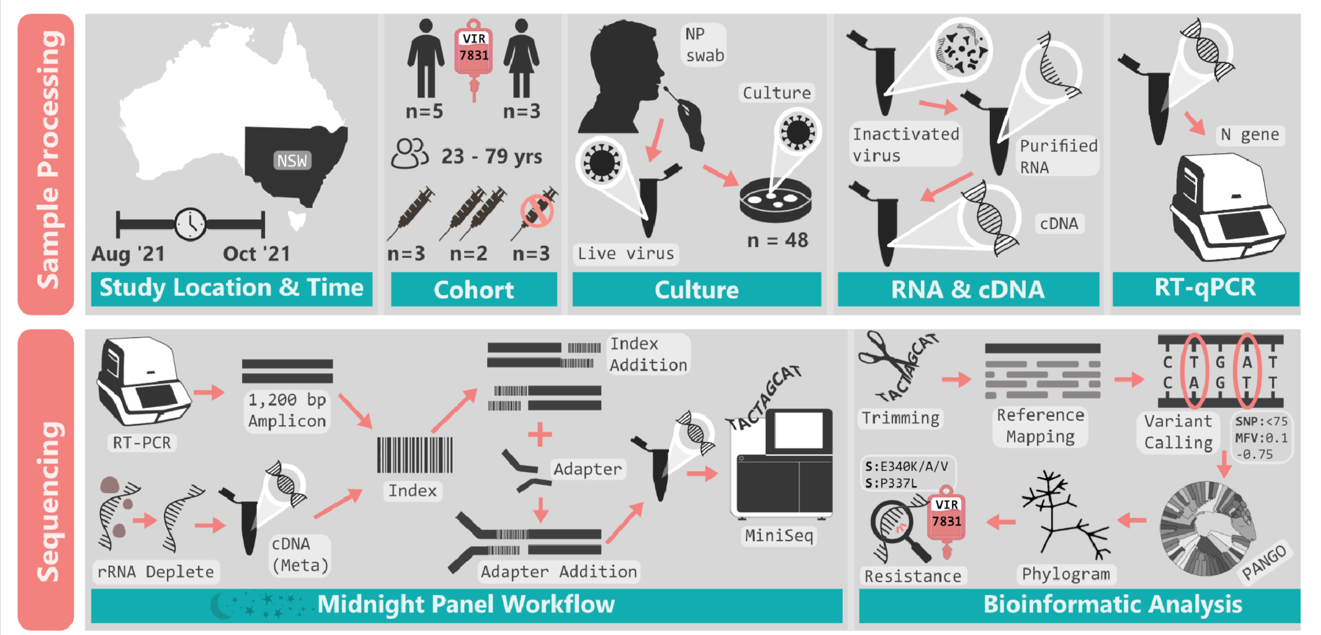


**Key:** NP swab – Nasopharyngeal swab; RT-PCR – reverse transcriptase real time polymerase chain reaction; N-gene – qRT-PCR amplifying the SARS-CoV-2 Nucelocapsid gene; rRNA deplete –rRNA Depletion (Human/Mouse/Rat), SNP – Single Nucelotide Polymorphism; MFV – Minority allele Frequency Variant; PANGO - Phylogenetic Assignment of Named Global Outbreak LINeages

**Figure S3.** Subsampled global phylogeny of SARS-CoV-2 highlighting Delta VOC genomes sequenced in this study

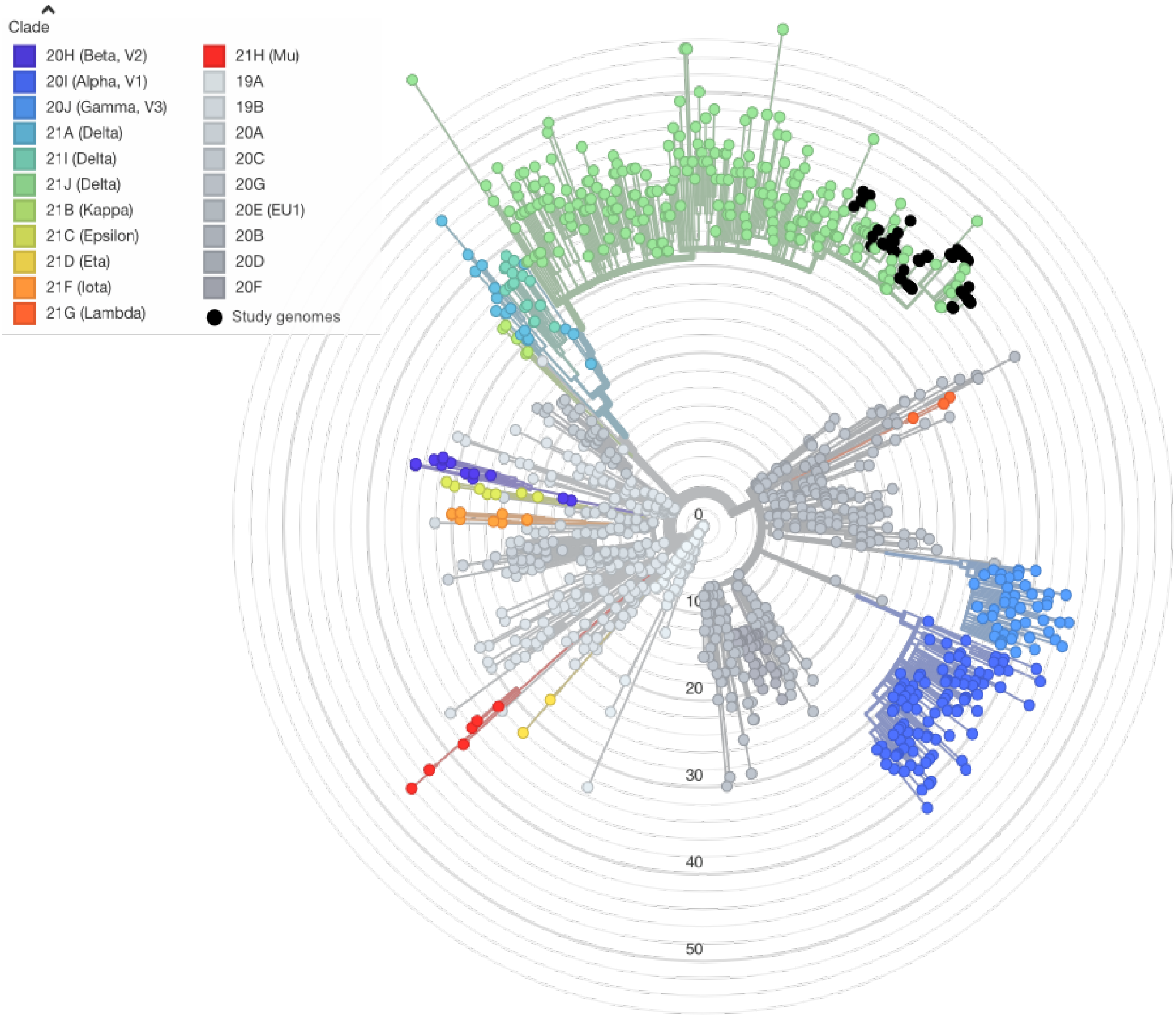


**Figure S3.** A maximum likelihood phylogency representing the global diversity of SARSCoV-2 genomes during the study period (n= 1,084). Genomes generated as part of this study are highlighted with back nodes and are part of the Delta VOC (GISAD clade 21J) (n=50). Node colours depict SARS-CoV-2 lineages circulating globally

**Table S2.** SARS-CoV-2 genomes containing consensus mutations at E340K in NSW, Australia

| Virus name | Accession ID | Lineage | COVID-19 treatments |
|---|---|---|---|
| hCoV-19/Australia/NSWICPMR-7006/2021 | EPI_ISL_4205292 | AY.39.1 | Sotrovimab |
| hCoV-19/Australia/NSWICPMR-9723/2021 | EPI_ISL_4968481 | AY.39.1 | Sotrovimab |
| hCoV-19/Australia/NSWSAVID-6825/2021 | EPI_ISL_5321824 | AY.39.1 | Unknown |
| hCoV-19/Australia/NSWSAVID-6814/2021 | EPI_ISL_5321822 | AY.39.1 | Unknown |

**Table S3.** SARS-CoV-2 genomes containing sub-consensus mutations conferring sotrovimab resistance in NSW, Australia

| Genome ID | Treatment | Conferring mutation (Read frequency) | GISAID Virus name |
|---|---|---|---|
| R013 | Sotrovimab | S:E340K (0.99) | Not uploaded (incomplete) |
| R017 | Unknown | S:E340K (0.99) | Not uploaded (incomplete) |
| R018 | Nil | S:E340K (0.99) | Not uploaded |
| R019 | Nil | S:P337T (0.99) | hCoV-19/Australia/NSW-ICPMR-11885 |
| R020 | Nil | S:P337L (0.11) | hCoV-19/Australia/NSW-NSW1877 |
| R021 | Nil | S:P337S (0.16) | hCoV-19/Australia/NSW-NSW4424 |

**Figure S4.** International incidence of S:E340K mutation: total 130 genomes

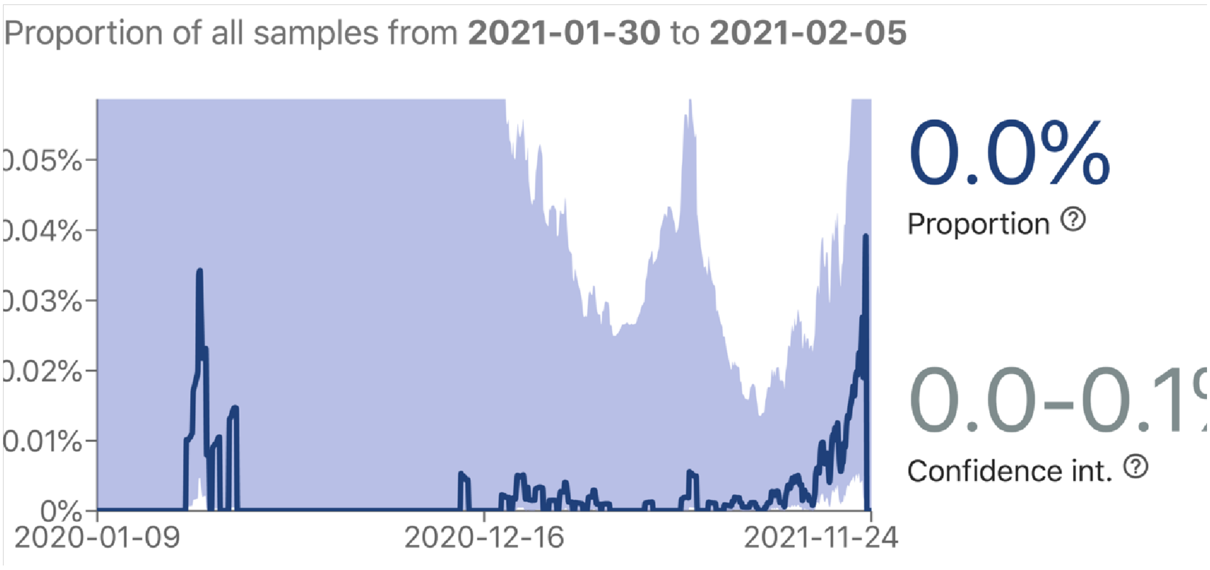

**Figure S5.** International incidence of S:E340A mutation: total 101 genomes

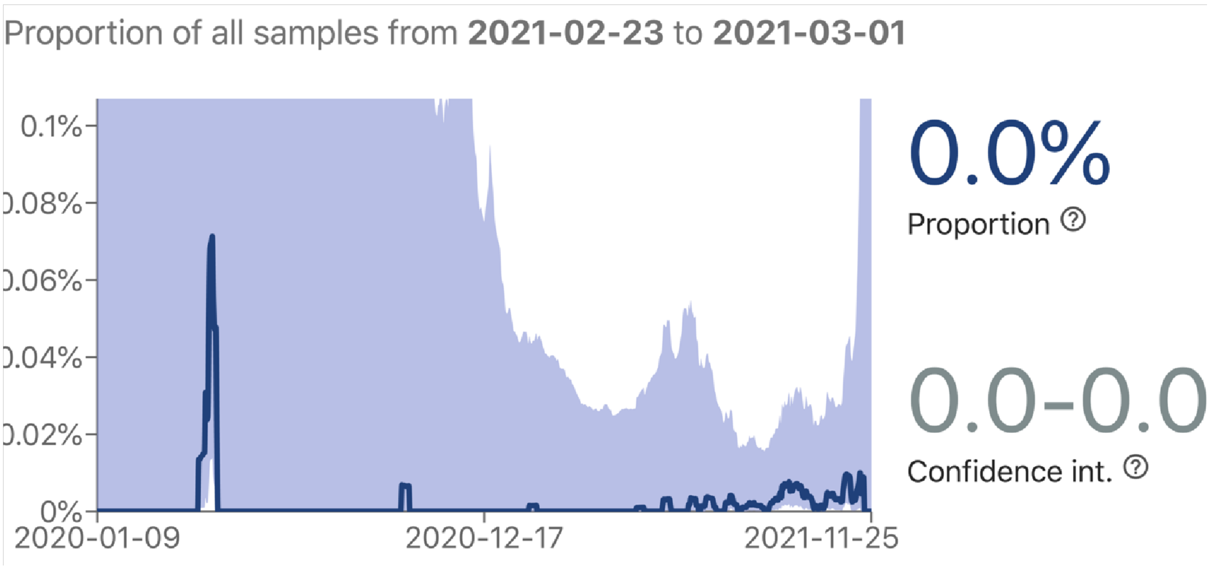


**Figure S6.** International incidence of S:E340V mutation: total 24 genomes

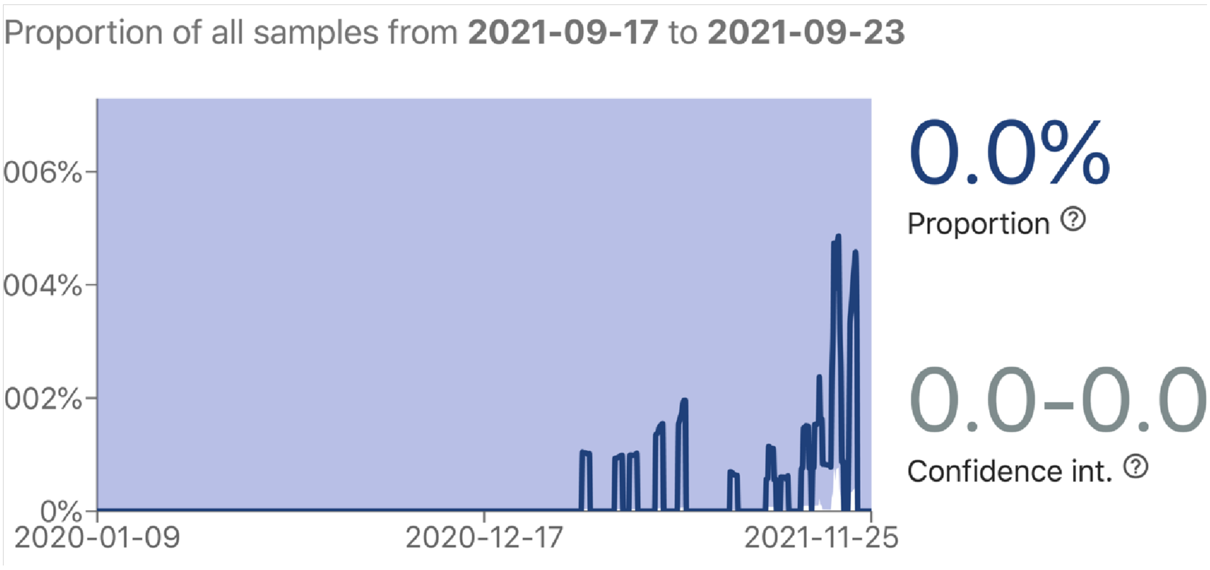

**Figure S7.** International incidence of P337L mutations: total 65 genomes

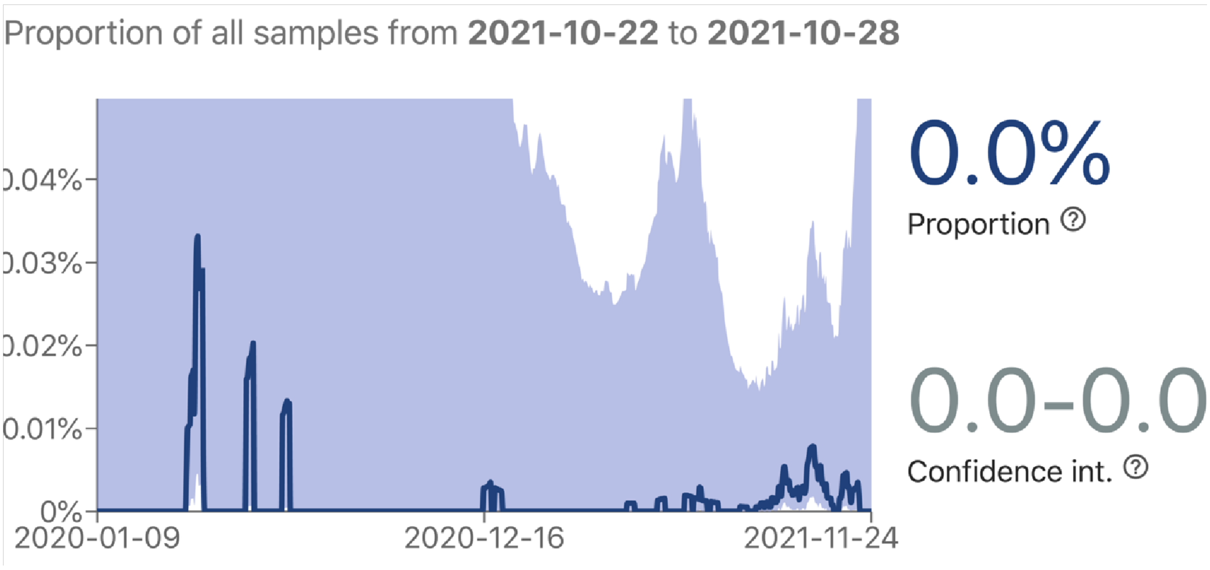
